# Personalized Immune Profiling in Pediatric Transplant Recipients: Linking Atypical B Cells to Vaccine Response

**DOI:** 10.1101/2025.03.25.25324615

**Authors:** Johannes Wedel, Ying Tang, Bayan Alsairafi, Vicki Do, Madeline Maslyar, Ryan Fleming, Marc A. Schwartz, Ulrike Gerdemann, Alexandre Albanese, Vanessa Mitsialis, Lauren V. Collen, Miki Nishitani, Mairead Bresnahan, Gwen Saccocia, Richelle Bearup, Ibeawuchi Okoroafor, Steven J. Siegel, Franziska Wachter, Katherine Waters, Nina Weichert-Leahey, Nigel J. Clarke, Kenneth D. Mandl, Leslie S. Kean, Scott B. Snapper, David M. Briscoe, Bruce H. Horwitz

## Abstract

Immunosuppression in solid organ transplant recipients inhibits protective immune responsiveness to pathogens and vaccines. However, specific cell states that associate with failure to generate protective immunity are not known. Here, we perform scRNAseq and CyTOF analyses of PBMC from 12 pediatric solid organ transplant recipients and 8 healthy children revealing the full spectrum of immune cell states present in these individuals, and examine the association of these cell states with the generation of protective humoral and cellular immunity following vaccination. We determined that clonal expansion of a subset of CD8^+^ effector T cells is significantly increased in immunosuppressed transplant recipients, and that increased frequencies of atypical B cells are associated with impaired humoral but intact T cell responses to vaccination. Interactome analysis suggests that robust cellular interactions between myeloid, T and B cells are required for successful protective immune responses to vaccination in pediatric transplant recipients.

**Graphical Abstract:** 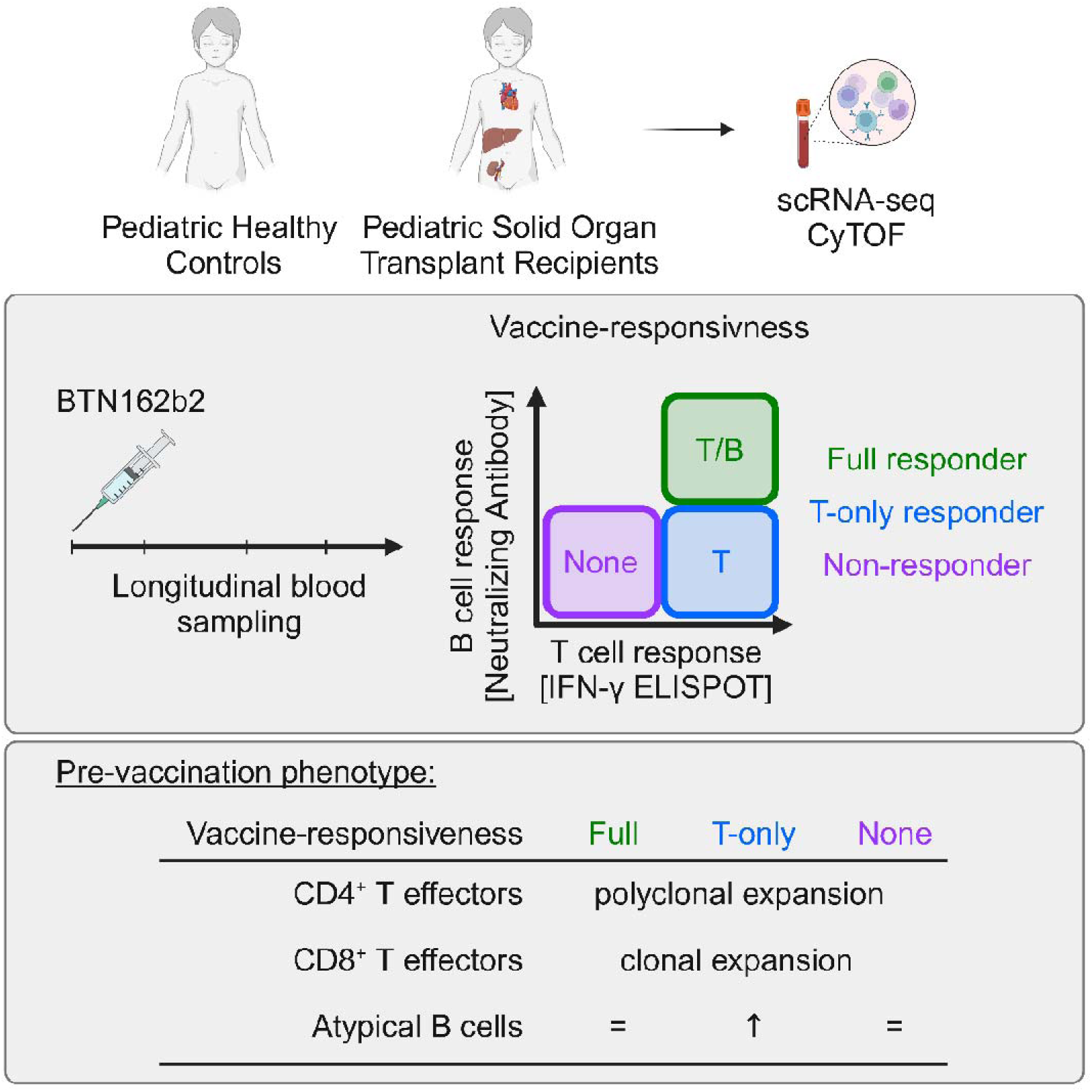

## Introduction

Organ transplantation is an effective treatment for children with end-stage organ failure, offering normal cognitive development, growth and quality of life. However, long-term graft survival following transplantation is dependent on lifelong treatment with immunosuppressive medications to prevent acute and/or chronic allograft rejection. These immunosuppressive medications can also inhibit protective immune responses necessary for defense against pathogens and response to vaccination. The risk inherent in this trade-off between immunosuppression necessary for graft survival and suppression of protective immune responses was most evident during the recent SARS-CoV-2 pandemic, which resulted in high morbidity and mortality among adult transplant recipients (1–4). Further, SARS-CoV-2 vaccine-induced responses were notably deficient in solid organ transplant recipients (SOTR) (5–7). To date, detailed studies determining whether immunologic defects in response to pathogens can be distinguished from immunosuppression required for graft survival are lacking but could potentially improve immunosuppressive strategies for SOTR.

In contrast to the adult population, morbidity and mortality among pediatric SOTR following SARS-CoV-2 infection were low, and complications were quite rare (8, 9). Nevertheless, responses to vaccination (anti-spike antibody production) were reported to be absent or blunted similarly in pediatric and adult transplant recipients (10). These results suggest that pediatric patients may generate alternative protective responses in the absence of specific antibodies, which could explain their positive outcomes following SARS-CoV-2 infection. It has been postulated that the generation of antigen-specific T cells following SARS-CoV-2 vaccination may provide a second protective layer in addition to the generation of Spike-specific antibodies. Indeed, there is compelling evidence for the generation of protective T cell responses in the absence of detectable antibodies against SARS-CoV-2 following immunization of nonhuman primates with Ad26.COV2.S (11). Nevertheless, little is known about the generation of antigen-specific T cell responses in pediatric SOTR following SARS-CoV-2 vaccination, nor whether antigen-specific T cell responses can be generated in the absence of vigorous humoral responses.

Here we provide a comprehensive analysis of peripheral immune cell states in pediatric SOTR and evaluate associations between specific cell subsets and humoral and cellular responses to SARS-CoV-2 vaccination. We have identified expansion of CD4^+^ and CD8^+^ effector T subsets and increased frequencies of atypical B cells in transplant recipients compared to healthy controls. In addition to identifying pediatric SOTR that exhibited fully intact humoral and antigen-specific T cells responses (Full responders) or those who lacked both of these responses (Non-responders) following vaccination, we identified a subset of pediatric SOTR who had defective production of neutralizing antibodies but robust antigen-specific T cell responses (T-only responders). The existence of T-only responders is consistent with the hypothesis that pediatric SOTR have the capacity to generate protective immune responses in the absence of successful antibody production. Interestingly, a significant increase in the frequency of atypical B cells was observed in T-only responders compared to healthy children. Further, integrating this dataset through interactome analysis suggested that specific interactions between CD4^+^, CD8^+^ and B cell subsets are important to sustain protective immunity. This study identifies specific cell subsets including clonally expanded effector CD8^+^ T cells that are found more frequently in pediatric SOTR than in healthy controls, and a T cell only response to SARS-CoV-2 vaccination in pediatric SOTR that is associated with the accumulation of atypical B cells. These results have important implications for the evaluation of successful immunosuppressive regimens in pediatric solid organ recipients.

## Results

### Immune profiling of stable pediatric solid organ transplant recipients

We used single cell RNA-sequencing (scRNAseq) and cytometry by time-of-flight (CyTOF) to comprehensively profile peripheral immune subsets in stable pediatric SOTR (Figure 1A). All SOTR were more than 9 months but less than 13 years post-transplant and were on maintenance immunosuppressant therapy. Twelve pediatric SOTR (n=9 kidney; n=2 liver; n=1 heart) aged between 5-19 years, and eight age-matched healthy controls (HC) were evaluated by scRNAseq (Table 1). A total of 169,741 cells passed quality control (Figure S1A-C) and were clustered into 5 immune cell lineages based on RNA lineage marker expression (Figure 1B-C and Figure S1D-E). Coarse T, B, and myeloid cell frequencies were similar in SOTR and HC; however, we found that SOTR had decreased frequencies of NK cells (7.2% *vs.*15.4%; *Q* = 0.03) and increased frequencies of plasma cells (8.8% *vs.* 3.9%; *Q* = 0.03) compared to healthy children (Figure 1D). Subclustering of NK/NKT and plasma cells in PBMC of SOTR *vs.* HC did not detect significant differences between SOTR and HC (Figure S2A-D). Within the myeloid compartment, we detected reduced numbers of dendritic subsets in SOTR compared to HC (Conventional dendritic cells: 4.6% *vs.* 10.2%, *Q* = 0.004; Monocyte-derived dendritic cells: 0.5% *vs.* 1.2%, *Q* = 0.02; Figure S2E-H), and trends toward expanded monocyte subsets (Figure S2G-H).

**Figure 1.**
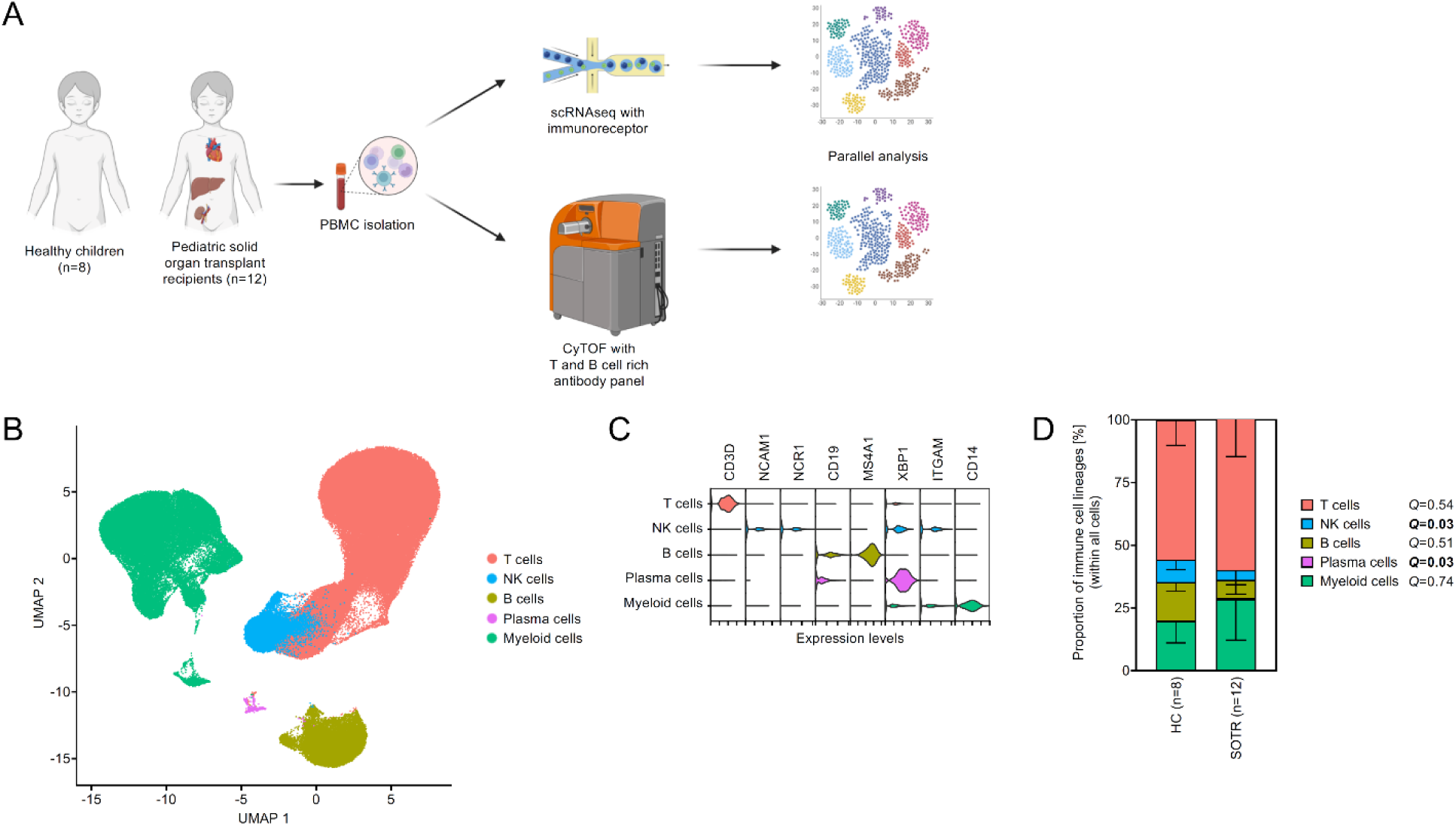
Single cell RNA-sequencing of PBMC from pediatric SOTR shows decreased frequencies of NK and increased frequencies of Plasma cells compared to healthy children. PBMC were isolated from 12 pediatric SOTR and 8 healthy children (HC) and subjected to scRNAseq including immunoreceptor sequencing (TCR/BCR), as well as CyTOF. **(A)** Experimental design. **(B)** UMAP of scRNAseq data depicts all cells clustered by immune cell lineage. **(C)** Violin plots illustrate markers that were used to identify immune cell lineages. **(D)** Stacked bar graph represents the percentage of each immune population within all cells per subject. Multiple testing-corrected *Q*-values of subset frequencies in SOTR *vs.* HC are given (mean ± SD; Mann-Whitney test with two-stage linear step-up procedure of Benjamini, Krieger and Yekutieli).

**Table 1.** Participants and demographics. *P* values were calculated using the Mann-Whitney test. *P* values <0.05 are highlighted in bold.

|  | HC (n=8) | SOTR (n=12) | *P* value |
| --- | --- | --- | --- |
| age [mean (min - max)] | 11 (5-13) | 12 (5-19) | 0.31 |
| sex [male: n (%)] | 3 (37.5%) | 7 (58.3%) | 0.65 |
| <u>Ethnicity:</u> |  |  |  |
| Hispanic or Latino [n (%)] | 0 (0%) | 7 (58.3%) | **0.01** |
| <u>Race:</u> |  |  |  |
| White/Caucasian [n (%)] | 8 (100%) | 7 (58.3%) | 0.11 |
| Black/African American [n (%)] | 0 (0%) | 0 (0%) | 1 |
| Asian [n (%)] | 0 (0%) | 1 (8.3%) | 1 |
| Other [n (%)] | 0 (0%) | 4 (33.3%) | 0.12 |

To examine quantitative and qualitative differences in T cell subsets between pediatric SOTR and HC, we subclustered CD8^+^ T cells into ten distinct subsets (Figure 2A). CD8^+^ subclusters were annotated based on the expression of markers associated with phenotypic naivety (*CCR7*, *S100A4*), activation status (cell surface *HLA-DR*, *LAG3* or transcription factors *TBX21*, *GATA3*, *EOMES*), effector molecules (*IFNG*, *GZMA*) and T cell receptor transcripts (*TRDC*, *TRGC*, *TRAV1-2*) (Figure 2B). We observed a significant expansion of one effector CD8^+^ T cell population (CD8^+^ T effectors 1) in transplant recipients compared to healthy controls (Figure 2C, D: 13.6% in SOTR *vs.* 5% in HC, *Q* = 0.004). Also, we found that CD8^+^ T cells from transplant recipients expressed significantly higher levels of proinflammatory transcripts, including *IFNG* and *CXCR3*, and activation-associated genes (HLAs, *TIGIT*, *LGALS1*, *STAT1*) than those in healthy controls (Figure 2E).

**Figure 2:**
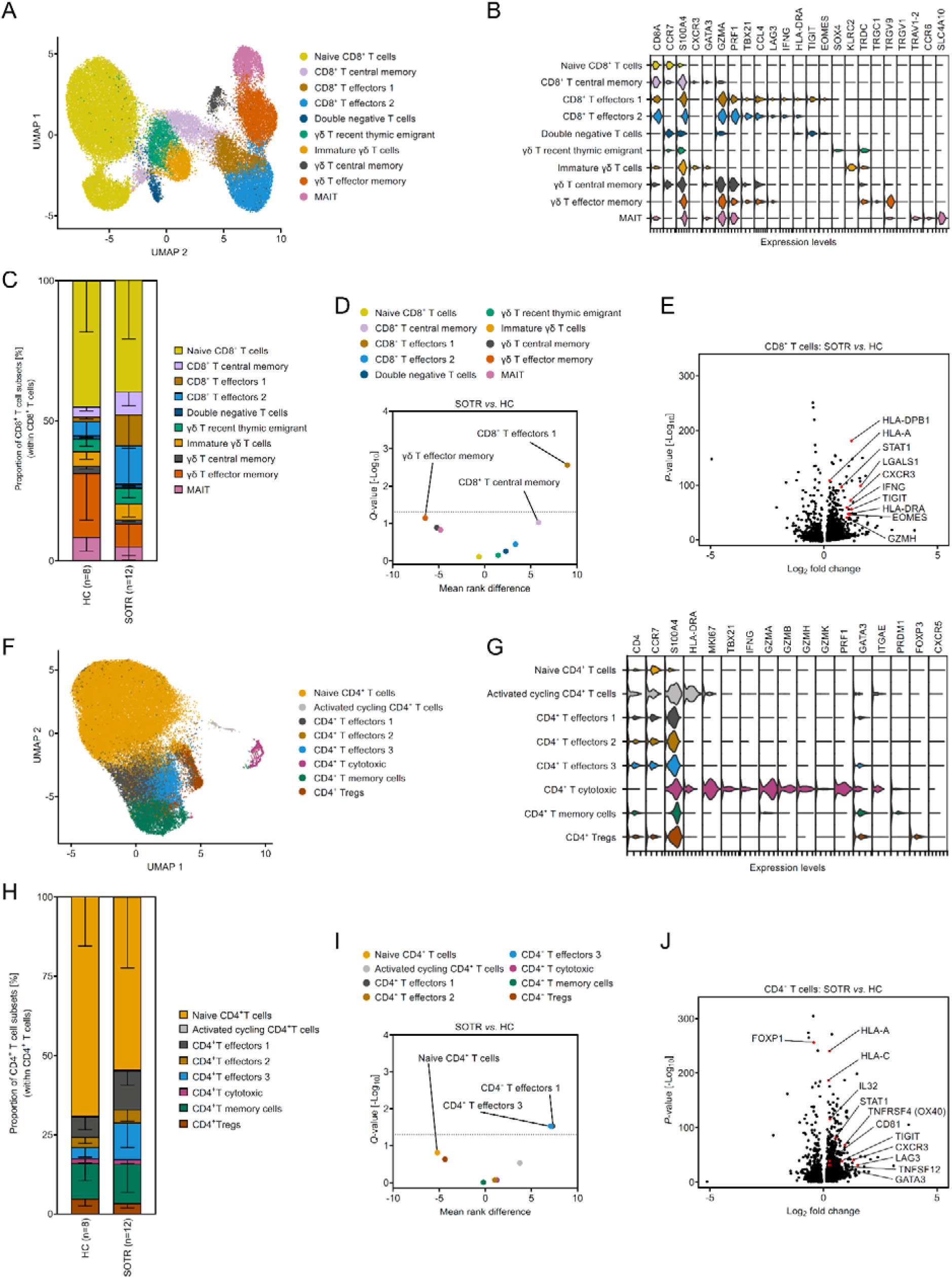
T cells from pediatric SOTR have increased frequencies of effector T cell subsets compared to healthy children. **(A-E)** CD8^+^ T cells were subsetted and reclustered to further analyze CD8^+^ T cell subset phenotypes. **(A)** UMAP of 46,793 CD8^+^ T cells from 20 participants (8 healthy controls and 12 SOTR) color-coded for CD8^+^ T cell subsets. **(B)** Violin plot illustrates markers used to identify each CD8^+^ T cell subset. **(C)** Stacked bar graph represents CD8^+^ T cell subcluster distribution per subject and cohort (mean ± SD). **(D)** Volcano plot depicts the mean rank difference and the corrected *Q*-value of CD8^+^ T cell subset frequencies in SOTR *vs.* HC (Mann-Whitney test with two-stage linear step-up procedure of Benjamini, Krieger and Yekutieli). **(E)** Volcano plot illustrates differential gene expression in CD8^+^ T cells in SOTR *vs.* HC. **(F-J)** CD4^+^ T cells were subsetted and reclustered to further identify CD4^+^ T cell subsets. **(F)** UMAP of 57,509 CD4^+^ cells from 20 participants (8 healthy controls and 12 SOTR) color coded for CD4^+^ T cell subsets. **(G)** Violin plot illustrates markers used to identify CD4^+^ T cell subsets. **(H)** Stacked bar graph represents CD4^+^ T cell subcluster distribution per subject and cohort (mean ± SD). **(I)** Volcano plot depicts the mean rank difference and the corrected *Q*-value of CD4^+^ T cell subset frequencies in SOTR *vs.* HC (Mann-Whitney test with two-stage linear step-up procedure of Benjamini, Krieger and Yekutieli). **(J)** Volcano plot illustrates differential gene expression in CD4^+^ T cells in SOTR *vs.* HC.

We also annotated CD4^+^ T cells based on markers of antigen experience, activation, transcription factors and effector molecules (Figure 2F-G). We identified eight distinct CD4^+^ T cell subsets, including expanded populations of effector CD4^+^ T cells in transplant recipients *vs.* HC (CD4^+^ T effectors 1: 12.1% *vs.* 6.5%, *Q* = 0.03; CD4^+^ T effectors 3: 11.6% *vs.* 3.5%, *Q* = 0.03; Figure 2H-I). CD4^+^ T cells from transplant recipients had higher levels of activation- and inflammation-associated transcripts including HLA molecules (*HLA-A*, *HLA-C*), *TNFRSF4* (OX40), *IL32*, *STAT1*, *CXCR3* (Figure 2J), similar to CD8^+^ T cells. Using pseudotime analysis, we grouped CD4^+^ T cells along a trajectory from naive to memory populations (Figure S3A) and evaluated the distribution of cells along this gradient. Transplant recipients had the highest abundance within effector subsets where HC had the highest abundance within naive subsets (Figure S3B). These findings suggest a relative shift in circulating CD4^+^ T cells subsets from naive to effector cells states following transplantation.

### TCR sequencing identifies clonal populations of effector CD8^+^ and CD4^+^ T cells in transplant recipients on immunosuppression

To compare the frequency of clonally related T cell subpopulations in PBMCs isolated from SOTR and HC, we integrated T cell receptor (TCR) sequencing data with the single-cell transcriptomic dataset for both CD4^+^ and CD8^+^ T cells (Figure 3). While both HC and SOTR had clonal populations within naive CD8^+^ T cells and mucosal-associated invariant T cell (MAIT) cells, SOTR showed a significant increase in clonally expanded populations of CD8^+^ T central memory, T effector, and γδ T effector memory subsets compared to HC (Figure 3A-B). This expansion was independent of the type of transplant, time post-transplant or immunosuppressive regimen (not shown). Furthermore, we observed that clonally expanded CD8^+^ T cells in SOTR were distributed across multiple effector subsets (Figure 3C). While we found some clonal expansion among CD4^+^ T cell subsets, it was much less pronounced in CD8^+^ T cells and was not significantly different between SOTR and HC (Figure 3D-F). Collectively, these data suggest that SOTR exhibit significantly higher levels of clonal CD8^+^ T cell expansion than healthy controls.

**Figure 3:**
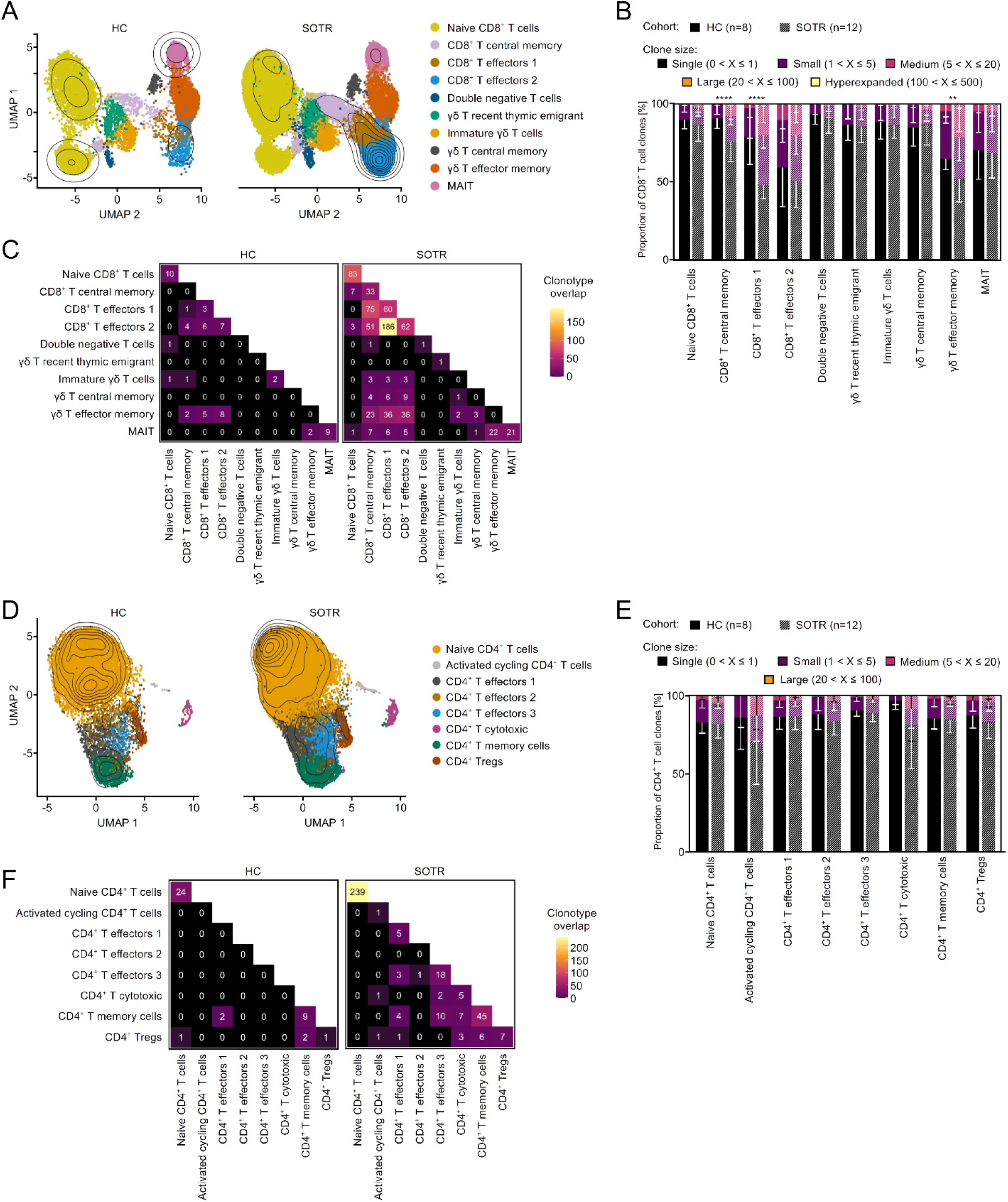
T effector cell expansion in SOTR is clonal in CD8^+^and polyclonal in CD4^+^ T effectors. **(A-C)** T cell receptor (TCR) sequences were mapped to CD8^+^ T cells. **(A)** Dots in UMAPs depict CD8^+^ T cell subsets from SOTR (right) or HC (left), and overlaid contour plots highlight the density of sequences identified ≥ 2 times (expanded clones). **(B)** Stacked bar graph depicts the proportion of clonally expanded CD8^+^ T cell subsets in SOTR (striped) and HC (filled) categorized by clonal size (Two-way ANOVA with Bonferroni correction; ** *P*<0.01, **** *P*<0.0001). **(C)** Heatmaps illustrate the number of CD8^+^ T cells that share identical TCR sequences across different CD8^+^ T cell subsets and within the same cluster in HC (left) and SOTR (right). **(D-F)** T cell receptor sequences were mapped to CD4^+^ T cells. **(D)** Dots in UMAPs depict CD4^+^ T cells from SOTR (right) or HC (left), and overlaid contour plots highlight the density of sequences identified ≥ 2 times (expanded clones). **(E)** Stacked bar graph illustrates the proportion of expanded CD4^+^ T cell subsets in SOTR (striped) and HC (filled) categorized by clonal size (Two-way ANOVA with Bonferroni correction; all comparisons are non-significant [*Padj*>0.05]). **(F)** Heatmaps illustrate the number of CD4^+^ T cells that share identical TCR sequences across different CD4^+^ T cell subsets and within the same cluster in HC (left) and SOTR (right).

### Expansion of a CD21^neg^ CD27^neg^ T-BET^pos^ CD11c^pos^ atypical B cell subset in pediatric transplant recipients

Turning towards B cells, we next subclustered circulating B cells into nine subsets (Figure 4A-B) and found a significant reduction in transitional B cells and marginal zone B cells in transplant recipients compared to healthy children (transitional B cells: 0.7% *vs.* 4.4%, *Q* < 0.0001; marginal Zone B cells 4.9% *vs.* 8.6%, *Q* = 0.05; Figure 4C-D). We also identified a significant increase in a subset of B cells characterized by high levels of *CD19*, *TBX21* and *ITGAX* expression, but low levels of *CR2* and *CD27* expression (6.4% vs. 1.5% in HC, *Q* = 0.03; Figure 4C-E), a phenotype aligns with previously reported atypical B cells or age-associated B cells (CD19^pos^ CD21^neg^ CD27^neg^ CD11c^pos^ T-BET^pos^) (12, 13). Consistent with the absence of allo-specific clonally expanded B cells within the blood, we found that the majority of B cell receptor (BCR) sequences, including those of atypical B cells, were diverse and only a small portion of BCRs (< 2%) were detected more than once (Figure 4F-H).

**Figure 4:**
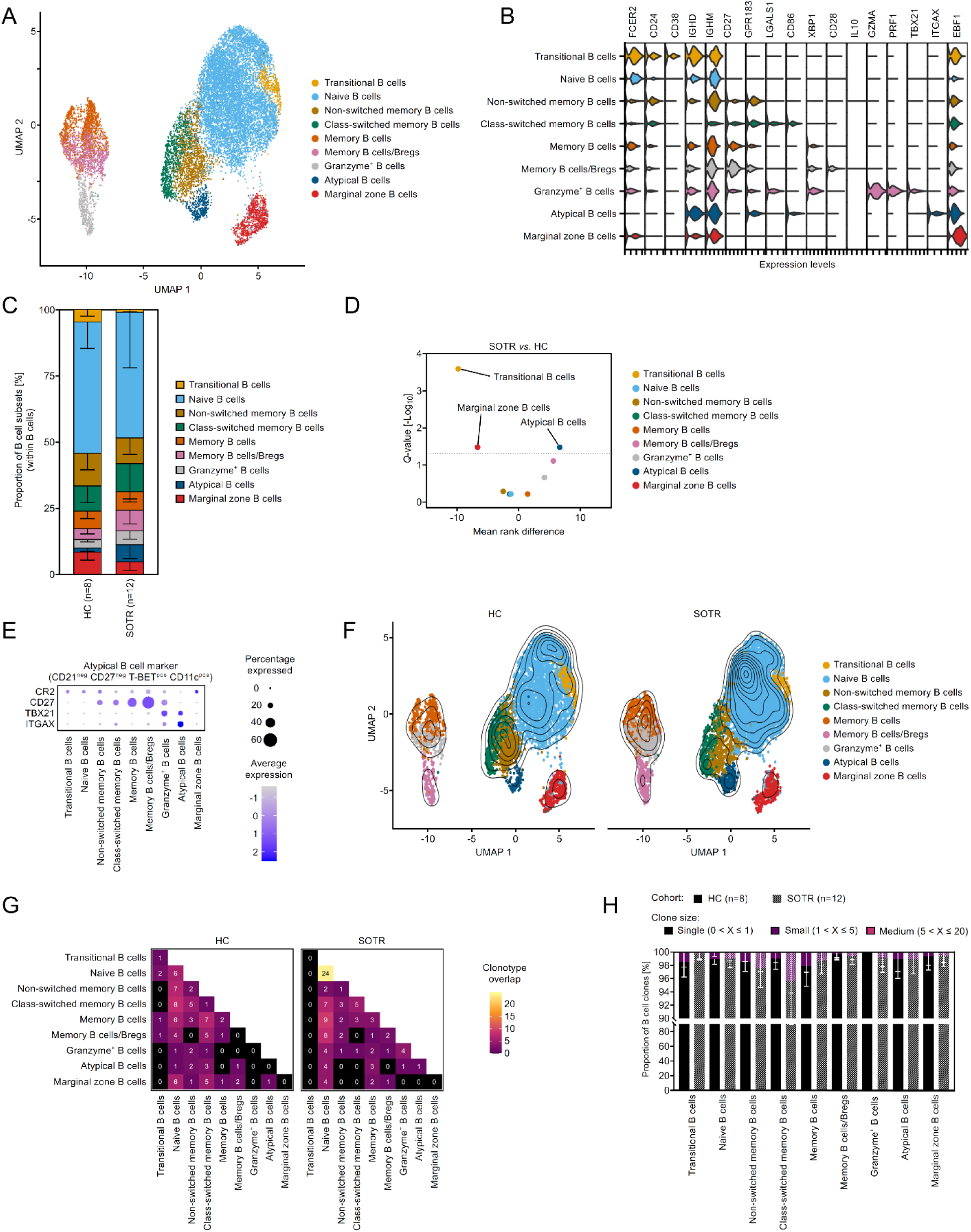
CD21^neg^ CD27^neg^ T-BET^pos^ CD11c^pos^ atypical B cells are expanded in pediatric SOTR compared to healthy children. B cells were subclustered and B cell subset phenotypes were evaluated. **(A)** UMAP of 16,383 B cells from 20 participants (8 healthy controls and 12 SOTR) color coded for B cell subsets. **(B)** Violin plot illustrates markers used to identify B cell subsets. **(C)** Stacked bar graph represents B cell subcluster distribution per cohort (mean ± SD). **(D)** Volcano plot depicts the mean rank difference and the corrected *Q*-value of B cell subset frequencies in SOTR *vs.* HC (Mann-Whitney test with two-stage linear step-up procedure of Benjamini, Krieger and Yekutieli). **(E)** Dot plot shows expression of markers used for the identification of the atypical B cell subset. **(F-H)** B cell receptor (BCR) sequences were matched with the transcriptomic data of B cells. **(F)** UMAPs depict B cells from HC (left) or SOTR (right). Identical BCR sequences that were detected ≥2 times (expanded B cell clones) are overlaid as the contour plot to illustrate clonal expansion of B cell subsets. **(G)** Heatmaps illustrate the number of B cells that share identical BCR sequences across different B cell subsets and within the same cluster in HC (left) and SOTR (right). **(H)** Stacked bar graph illustrates the proportion of expanded B cell subsets in SOTR (striped) and HC (filled) categorized by clonal size (Two-way ANOVA with Bonferroni correction; all comparisons are non-significant [*Padj*>0.05]).

To further explore alterations in immune cell populations within the peripheral blood of SOTR and confirm our transcriptomic findings, we performed CyTOF analysis of PBMCs using a T and B cell marker rich antibody panel (Table S1). Similar to the scRNAseq data, we did not observe significant changes within major immune cell lineage frequencies (Figure S5A-E). There was a trend towards lower frequencies of naive T cells and higher frequencies of effector CD4^+^ and CD8^+^ T cells in SOTR compared to HC, but these differences were not statistically significant (Figure S5F-H). Interestingly, when looking into the B cell subclusters in CyTOF data, we identified two atypical B cell subsets: one expressed IgM and IgD, while the other lacked both, suggesting that this latter population had already undergone class switching from surface expression to secretion (Figure 5A-B). Both atypical B cell subsets were significantly increased in SOTR *vs.* HC (IgM^pos^/IgD^pos^: 7.6% *vs.* 1.2%, *Q* = 0.003; IgM^neg^/IgD^neg^: 2.1% *vs.* 0.5%, *Q* = 0.04, Figure 5C-D). Collectively, CyTOF and scRNAseq data showed remarkably similar trends regarding expanded effector/memory T cell subsets and the expansion of atypical B cells in SOTR (Figure S5I).

**Figure 5:**
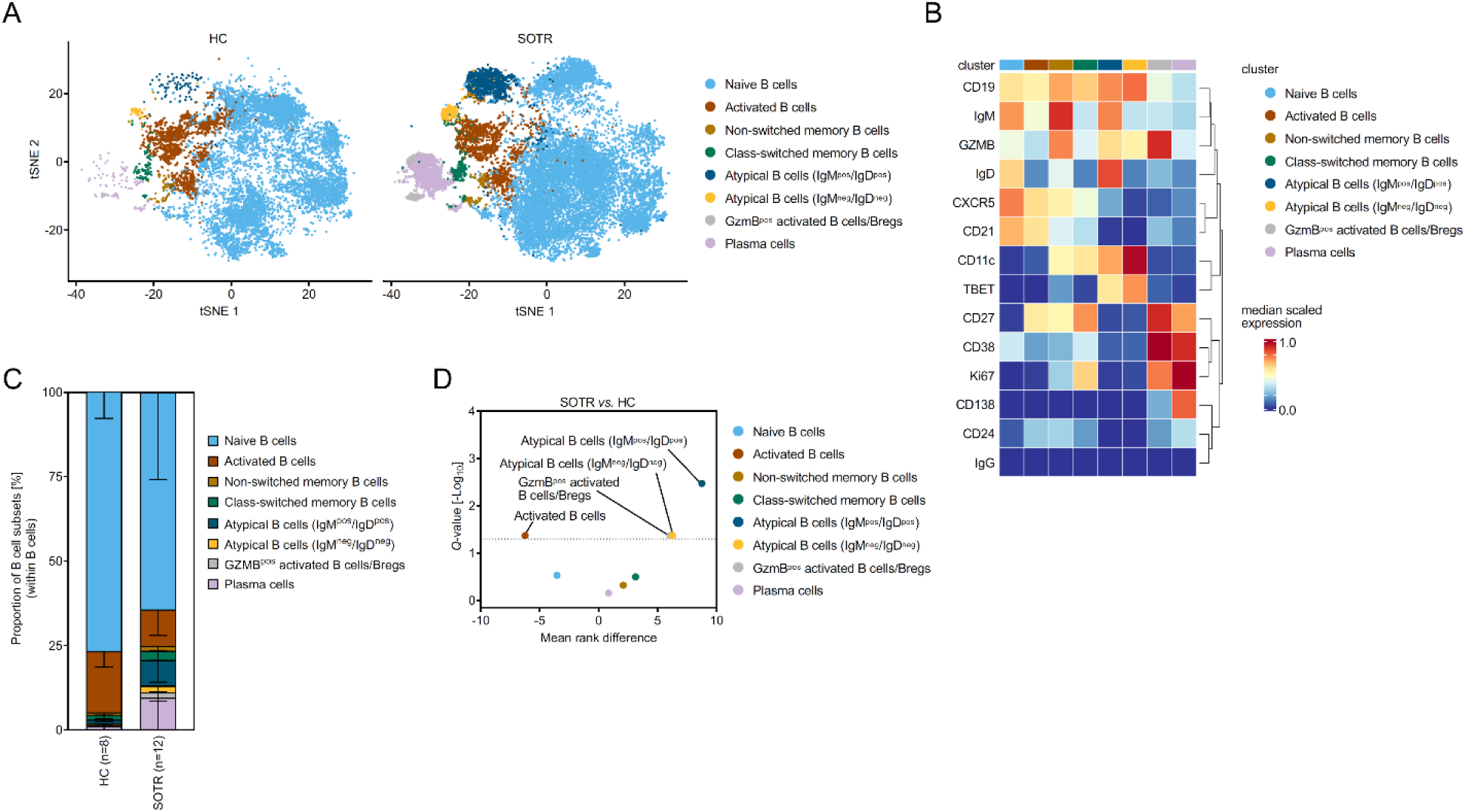
Atypical B cells can be divided into IgD^neg^/IgM^neg^ and IgD^pos^/IgM^pos^ subsets and both are expanded in pediatric SOTR compared to HC. PBMC samples from all 12 pediatric SOTR and 8 healthy children (HC) were analyzed using CyTOF (see also Figure S5). B cells were reclustered to identify and quantify B cell subsets. **(A)** tSNE plots depict B cell subsets split by healthy controls (left) and SOTR (right). **(B)** Heatmap illustrates B cell marker expression that was used to annotate B cell subsets. **(C)** Stacked bar graph represents B cell cluster distribution in SOTR and HC (mean ± SD). **(D)** Volcano plot depicts the mean rank difference and the corrected *Q*-value of B cell subset frequencies in SOTR *vs.* HC (Mann-Whitney test with two-stage linear step-up procedure of Benjamini, Krieger and Yekutieli).

### Association between immune profiles and protective immune responses

We postulated that specific immune profiles identified in our transplant subjects might be linked to altered immune responses to vaccination. To evaluate this possibility, we compared the humoral and cellular responses of SOTR and HC following administration of the SARS-CoV-2 BNT162b2 mRNA vaccine. Plasma and PBMC samples were collected from all subjects (SOTR and HC characterized above) pre- and one week post-initial and second BNT162b2 immunizations, and at 12 weeks post initial vaccination (Figure 6A). Some transplant recipients received a third BNT162b2 booster vaccination at 12 week post initial vaccination in which case we collected additional samples 1- and 3-weeks post booster vaccination (Figure 6A). All enrolled subjects were initially negative for ancestral nucleocapsid protein, indicating no prior exposure to SARS-CoV-2 virus (data not shown). Selected examples of antibody responses against ancestral S1, S2 and RBD to vaccination over time in both HC and SOTR are shown in Figure S5A-B. We found that while all HC (n=8) generated neutralizing spike antibodies to ancestral and delta RBD when measured one week after the second vaccination, only 3 out of 12 SOTR achieved over 50% neutralization (Figure 6B). All HC and two of these three SOTR had >50% neutralization of omicron RBD interactions with ACE2. In contrast, one SOTR exhibited a lower neutralization level of 36%, potentially indicating a limited and oligoclonal humoral response (Figure 6B; right panel). We also evaluated vaccine-specific CD4^+^ T cell-dependent immunity following BNT162b2 administration by measuring spike-induced IFNγ secretion using ELISPOT (Figure 6C-D). Spike protein-specific CD4^+^ T cell responses were observed in all HC (data not shown) as well as in 3 out of 3 transplant recipients with >50% antibody mediated RBD neutralization (designated as Full responders). Interestingly, while 4 out of the 9 patients who failed to generate >50% neutralizing antibody also failed to develop spike-specific CD4^+^ T cell responses (designated as Non-responders), the remaining 5 exhibited spike protein-specific IFNγ production by CD4^+^ T cells (designated as T-only responders) (Figure 6D-E).

**Figure 6:**
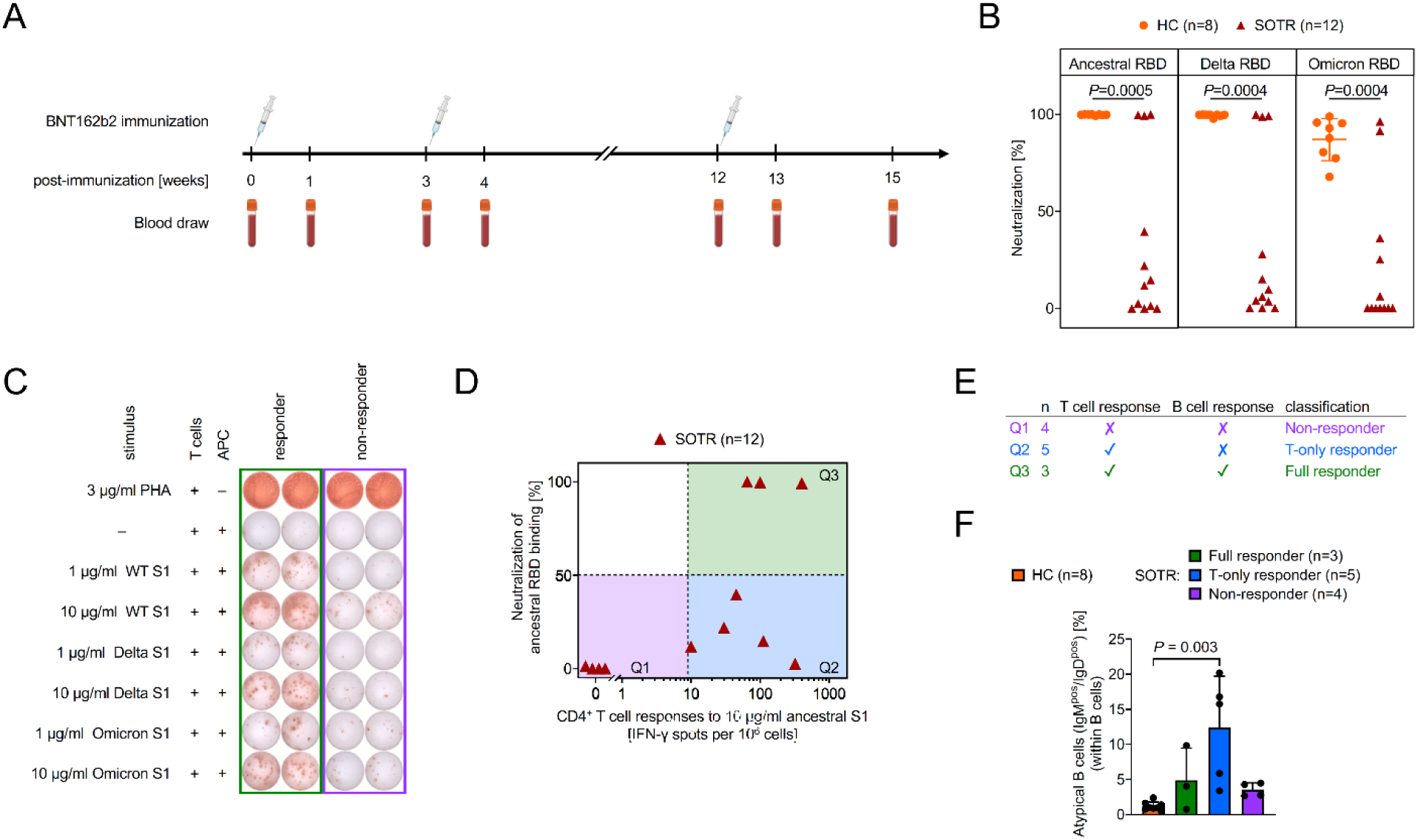
Increased frequencies of IgM^pos^/IgD^pos^ atypical B cells in pediatric solid organ transplant recipients are associated with the development of intact T cell-but impaired humoral responses following BNT162b2 immunization. Plasma and PBMC samples were obtained prior and 1 week post immunization with first and second doses of BNT162b2, and either at 3 months following 1^st^ immunization or prior and 1- and 3-weeks post 3^rd^ immunization (booster dose). **(A)** Schematic of the timeline. **(B)** Neutralizing-antibody concentrations against the RBD of ancestral, B.1.617.2 (delta) and B.1.1.529 (omicron) SARS-CoV-2 variants following the second vaccination. **(C-D)** Frequency and function of spike-specific CD4^+^ T cell responses were assessed by IFN-γ ELISPOT one week following the second immunization. Stimulation of T cells with phytohaemagglutinin (PHA, 3 µg/ml) was used as a technical positive control. **(C)** Representative image of the IFN-γ ELISPOT assay depicting a responder and a non-responder to BNT162b2 vaccination. **(D)** Ancestral S1-specific CD4^+^ T cell activation is compared to neutralizing-antibody concentrations (depicted in Panel B) in pediatric SOTR. **(E)** SOTR were categorized into three groups based on humoral and cellular responses as Non-responders (Q1; no T and no humoral responses), T-only responders (Q2; normal T cell responses [≥ 10 IFN-γ spots/10^6^ cells] but defective B cell responses [<50% neutralization]) and Full responders (Q3; T and humoral responses). **(F)** Bar graph compares frequencies of IgM^pos^/IgD^pos^ atypical B cells between HC and SOTR subgroups (Kruskal-Wallis test with Dunn’s multiple comparison).

We next investigated whether specific alterations in immune cell subset frequencies were associated with any of these 3 classes of responders (Figure S5C-E). We found a significant increase in the frequency of IgD^pos^/IgM^pos^ atypical B cells identified by CyTOF in T-only responders compared to HC (Figure 6F). In contrast, we did not detect a significant difference in the frequencies of IgD^neg^/IgM^neg^ atypical B cells between groups (Figure S5F). By scRNAseq, the frequencies of Immature γδ T cells were significantly reduced in T-only responders compared to Non-responders (*P*=0.02, Figure S5G). Regarding immunosuppression, we noted a significant higher use of sirolimus-based regimens in T-only responders (Figure S5D and Table 2; *P*=0.03). Impaired responsiveness (T-only and Non-responders combined) correlated with greater use of MMF-based immunosuppression (Figure S5E), but this did not reach statistical significance (Table 2). Collectively, these comprehensive immune profiling data demonstrate that while most transplant recipients have the potential to mount antigen-specific T cell responses following BNT162b2 vaccination, T-only responders with elevated levels of atypical B cell are less likely to simultaneously mount neutralizing B cell responses.

**Table 2.** Demographics of pediatric SOTR grouped by vaccine responsiveness. *P* values were calculated using the Kruskal-Wallis test. *P* values <0.05 are highlighted in bold.

|  | Full responder<br>(n=3) | T-only responder<br>(n=5) | Non-responder<br>(n=4) | *P* value |
| --- | --- | --- | --- | --- |
| age [mean (min - max)] | 13 (11 - 15) | 13.8 (8 - 19) | 8.5 (5 - 14) | 0.21 |
| sex (male (%)) | 2 (33.3%) | 1 (20%) | 4 (100%) | 0.07 |
| BMI [mean (min - max)] | 20.4 (17.7 - 23.0) | 19.0 (14.4 - 24.4) | 17.4 (13.9 - 20.8) | 0.77 |
| <u>Ethnicity:</u> |  |  |  |  |
| Hispanic or Latino [n (%)] | 1 (33.3%) | 2 (40%) | 4 (100%) | 0.15 |
| Not Hispanic or Latino [n (%)] | 2 (66.7%) | 3 (60%) | 0 (0%) | 0.15 |
| <u>Race:</u> |  |  |  |  |
| White/Caucasian [n (%)] | 3 (100%) | 3 (60%) | 1 (25%) | 0.20 |
| Black/African American [n (%)] | 0 (0%) | 0 (0%) | 0 (0%) | 1 |
| Asian [n (%)] | 0 (0%) | 1 (20%) | 0 (0%) | 1 |
| Other [n (%)] | 0 (0%) | 1 (20%) | 3 (75%) | 0.09 |
| <u>Organ:</u> |  |  |  |  |
| kidney (n=9) | 2 (66.7%) | 4 (80%) | 3 (75%) | 1 |
| liver (n=2) | 1 (33.3%) | 0 (0%) | 1 (25%) | 0.47 |
| heart (n=1) | 0 (0%) | 1 (20%) | 0 (0%) | 1 |
| years post-transplant [mean (min - max)] | 9.3 (4.5 - 12.3) | 6.5 (5 - 8.6) | 3.7 (0.8 - 9) | 0.19 |
| HLA class I mismatch [mean (min - max)] | 5 (4-6) | 2.5 (1-4) | 4.3 (3-5) | 0.13 |
| HLA class II mismatch [mean (min - max)] | 2.5 (2-3) | 3 (1-5) | 2.3 (1-3) | 0.9 |
| <u>immunosuppressive drugs:</u> |  |  |  |  |
| tacrolimus [n (%)] | 3 (100%) | 1 (20%) | 3 (75%) | 0.12 |
| azathioprine [n (%)] | 2 (66.7%) | 0 (0%) | 2 (50%) | 0.13 |
| sirolimus [n (%)] | 0 (0%) | 4 (80%) | 0 (0%) | **0.03** |
| MMF [n (%)] | 0 (0%) | 4 (80%) | 2 (50%) | 0.13 |
| prednisolone [n (%)] | 0 (0%) | 1 (20%) | 1 (25%) | 1 |
| belatacept [n (%)] | 0 (0%) | 1 (20%) | 0 (0%) | 1 |

### Protective immune responses require interactions among myeloid, CD4^+^, CD8^+^ and B cell subsets

Finally, we employed an interactome analysis of our scRNA-seq dataset in order to determine the mechanism of responsiveness among all immune subsets following transplantation. Despite the treatment with immunosuppression, we observed increased cell-cell interactions between each cell lineage in SOTR compared to healthy controls (Figure 7A) suggesting an increased level of immune activation post-transplantation. Nevertheless, while the number of interactions between myeloid, CD4^+^, CD8^+^ and B cells was reduced in T-only responders and Non-responders compared to Full responders, reduction of interactions among T and B cells was most notable (Figure 7B). These findings suggest that protective immunity relies on coordinated interactions among myeloid, T, and B cell subsets, highlighting that distinct communication networks are required for mounting effective cellular and/or humoral responses following vaccination. Evaluating these signals as part of a personalized immune profiling approach could serve as a valuable tool to improve immunosuppressive therapy in pediatric SOTR by optimizing protective immune responses following vaccination while preserving graft function and survival.

**Figure 7:**
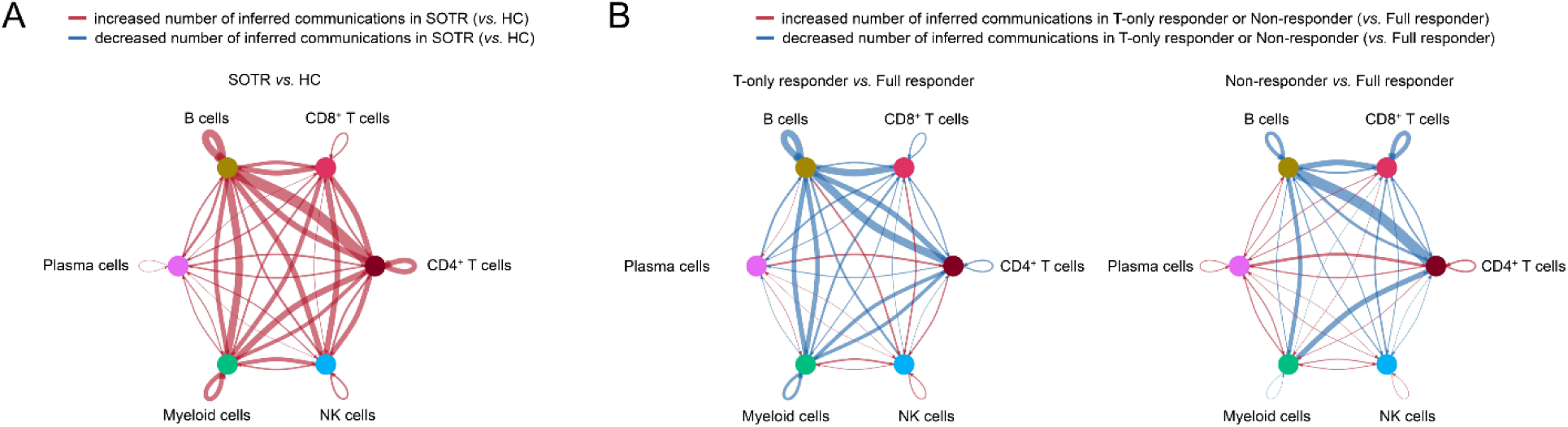
Protective immune responses require interactions among myeloid, CD4^+^, CD8^+^ and B cell subsets. **(A)** Circle plot shows the relative change of inferred intercellular interactions within immune cells between SOTR and HC. **(B)** Circle plots show the relative change of inferred intercellular interactions within immune cells between T-only responder (left) or Non-responder (right) compared to Full responder. The color represents either an increase (red) or decrease (blue) and the width represents the number of differential interactions.

## Discussion

This study provides a comprehensive single-cell analysis of the immune landscape in pediatric SOTR. SOTR exhibit expansion of CD4^+^ and CD8^+^ T effector subsets, which is oligoclonal for CD8^+^ and polyclonal for CD4^+^ T cells. Notably, a subgroup of SOTR had increased frequencies of atypical B cells, which were associated with impaired B cell but intact T cell responses following vaccination. There were no major differences in the frequencies of T or innate cell subsets between SOTR who mount antibody responses and those who lacked T and/or B cell responsiveness to BNT162b2 vaccination. However, interactome analysis suggests that impaired vaccine responses are associated with reduced interactions between myeloid, CD4^+^, CD8^+^ and B cells in SOTR.

One of the key insights from our study is the differential impact of immunosuppression on CD4^+^ and CD8^+^ T cell subsets. While both CD4^+^ and CD8^+^ T cells had shifts from naive to activated effector subsets in SOTR, the CD8^+^ T effector expansion was oligoclonal, contrasting to a polyclonal expansion of CD4^+^ T effector subsets. This suggests that current immunosuppressive regimens effectively target and inhibit alloimmunity in CD4^+^ T cells but are less effective in suppressing alloimmune CD8^+^ T cell responses. In addition, CD8^+^ clones were distributed across multiple effector subsets suggesting either that individual clones expand and differentiate into distinct subsets or that some related subsets represent a transitional cell state(s) towards fully differentiated effector cells. Nevertheless, the persistent expansion of effector CD8^+^ T cell clones in SOTR compared to healthy controls indicates potential alloreactivity, although increased responses to other antigens, such as pathogens, are also possible. If alloreactivity is indeed at the root of CD8^+^ T cell activation, our results raise the question of whether improved CD8^+^ targeted immunoregulatory therapy could improve graft survival.

We expected that the phenotype and/or subset composition of immune cell subsets in SOTR with impaired vaccination responses would be different from that observed in Full responders. Indeed, we identified an expansion of atypical B cells in a subset of SOTR that had intact T cell-but lacked B cell responsiveness following vaccination. Despite observing their association with impaired immunization-induced antibody production, our study was observational and therefore unable to definitively determine why atypical B cells are linked to impaired antibody production following vaccination. Increased frequencies of atypical B cells have been observed in several chronic inflammatory conditions (14–16) and they share transcriptional profiles across different immune-mediated diseases (17–19). These cells are thought to be antigen-experienced cells that develop in type I proinflammatory environments (13). Additionally, it has been reported that atypical B cells might expand in the absence of exogenous antigen in chronic inflammatory conditions and could be autoreactive or recognize autoantigens (20). In regard to solid organ transplantation, atypical B cells have not been well characterized. Brinas *et al.* found an expansion of a CD19^pos^CD11c^pos^CD27^neg^ B cell subset in the blood of adult kidney transplant recipients more than 5 years post-transplantation compared to recipients within the first year post-transplant or healthy volunteers (21). Importantly, this expansion was not associated with the chronological age of the SOTR but with the time post-transplantation (21). As transplant rejection is a type I proinflammatory process, it is plausible that atypical B cells expand in the absence of antigen and their function extends beyond antibody secretion to include regulation of B cell-dependent immunity (22). It is also possible that SOTR with higher frequencies of atypical B cells not only show impaired B cell responsiveness to vaccination but also to alloantigen. Consistent with this hypothesis, in the study by Brinas *et al.* (21), all kidney transplant recipients with increased atypical B cells tested negative for donor-specific antibodies (DSA), while DSA positivity in recipients with low frequencies of atypical B cell was 11%. Mechanistic studies focusing on the role of atypical B cells in transplantation are needed to elucidate their impact on protective *vs.* allospecific immune function and evaluate if atypical B cells can serve as a biomarker to identify SOTR with low risk for antibody mediated rejection.

The differential impact of immunosuppressive regimens in SOTR might be another relevant factor on vaccine responsiveness. We noted that Full responders primarily received a regimen of tacrolimus and azathioprine, whereas T-only responders were on sirolimus and MMF therapy. Non-responders had a combination of tacrolimus with either azathioprine or MMF. Notably, only the use of sirolimus appeared to be significantly different among the three groups. Indeed, *in vitro* experiments suggest that tacrolimus may spare the humoral response to some degree, whereas sirolimus has direct inhibitory effects on vaccination-induced B cell activation and differentiation while allowing T cell responses (23). Consistent with our trends in pediatric SOTR, other studies in adults have shown that tacrolimus/MMF-based immunosuppression is associated with impaired seroconversion and impaired T follicular helper responsiveness (24–27). However, this observation may be specific to SOTR as we did not find any impaired seroconversion in pediatric bone marrow transplant recipients or in immunosuppressed children with inflammatory bowel disease (data not shown).

Finally, it remains unclear whether isolated T cell immunity in T-only responders provides adequate protection from transmission and/or severity of SARS-CoV-2 disease. In non-human primates, multiple studies showed that T cell immunity is important for antiviral control and viral clearance (11, 28, 29). For example, in a dose-down study, T cell responses, neutralizing and binding antibody titers were preserved while Fc receptor binding and functional antibody levels were impaired at low vaccine doses (11). In subsequent challenge experiments, neutralizing antibody and Fc receptor binding titers were linked to the prevention of transmission, and T cell immunity was identified to be important for antiviral control (11). These data suggest that T-only responders are potentially not protected from transmission of SARS-CoV-2 but could be protected from severe disease following vaccination.

Our study has two major limitations: First, the small sample size is a limiting factor. Nevertheless, this study is one of the largest characterizations of the immune landscape in immunocompromised pediatric SOTR that links immune cell phenotypes to functional vaccination responsiveness. This study used the unique opportunity of BNT162b2 immunization during the SARS-CoV-2 pandemic to characterize a truly naive protective immune response in SARS-CoV-2 inexperienced pediatric transplant recipients. During enrollment, many pediatric patients received remote care to minimize the risk of exposure to a potentially harmful infection and we were unable to recruit more individuals. However, we were able to also include healthy children as a control group and identify transplant-associated changes to complement other profiling studies such as by Rao et al. (30). A second limitation is the absence of a second independent validation cohort which is also due to the limited numbers of enrolled patients. Both limitations will be subject to future investigations in this area.

In conclusion, our study underscores the differential impact of immunosuppression on CD4^+^ and CD8^+^ T cells in pediatric SOTR and highlights the need for improved therapies to better control CD8^+^ T cell responses. The association of atypical B cells with impaired vaccination responses emphasize the importance of further research to elucidate the mechanisms underlying immune dysregulation in pediatric SOTR and their relevance for alloimmunity. Our findings provide a foundation for future studies aimed at optimizing immunosuppressive regimens using personalized immunoprofiling and improving immune function in this vulnerable population.

## Methods

### Sex as a biological variable

Our study examined male and female study participants, and similar findings are reported for both sexes.

### Patient selection and sample collection

Pediatric solid organ transplant recipients (age 5-19 years) followed in ambulatory clinics at Boston Children’s Hospital and age-matched healthy volunteers were prospectively enrolled in this study (Table 1). After providing written informed consent, whole blood was collected in K2 EDTA tubes pre- and 7 days post BNT162b2 vaccination and either at 90 days post-enrollment (2 doses of BNT162b2) or 21 days post third BNT162b2 vaccination. Plasma was separated and stored at −80^°^C prior to use. PBMC were isolated using high density-gradient centrifugation (GE Healthcare, Chicago, IL) and cryopreserved in 10% DMSO/FBS until analysis.

### Single cell RNA-sequencing

PBMC were thawed for bulk analysis in two independent library preparations/sequencing runs. Cells were stained with hashtag antibodies (Table S2) in 1% BSA in PBS for 30 min at room temperature, washed twice in 1% BSA in PBS and cell number was assessed by grid counting. Cell viability was consistently >80%. Equal proportions of cells from each sample were pooled (16-20 samples/library preparation) and 40k pooled cells were loaded onto each of sixteen Chromium Next GEM Single Cell 5’ Kit v2 channels (Illumina, San Diego, CA). Library preparations were performed according to manufacturer’s instruction and libraries for gene expression, immunoreceptors, and hashtag-antibodies were pooled and sequenced on a Novaseq 6000 S4 (Illumina, San Diego, CA). Raw reads were aligned to the human GRCh38 genome using Cell Ranger pipeline (version 7.0.1) (31) and cells that passed QC (nFeature_RNA > 400, nCount_RNA > 1250, percent.mt < 10, nCount_HTO < 4000) were imported into a Seurat object (R version 4.3.2, Seurat version 5.0.1) (32). Samples were demultiplexed using the hashtag signal (99.9 percentile) and cells with multiple hashtag signals were considered doublets and excluded from analysis. Additionally, TCR and BCR genes were excluded from transcriptomic analysis to reduce variation. Both independent library preparations were integrated using harmony (version 1.2.0) (33) and cluster resolution was determined using the Clustree method (version 0.5.1) (34). Cell annotation was performed using curated marker gene lists extracted from reference atlases (35), and compared to ScType (version June 2021) (36) and Azimuth (version 0.4.6) (37) annotation tools (Figure S1E and data not shown). Transcriptionally indistinguishable clusters were manually merged. Differential transcript expression was performed using the DESeq2 method (version 1.40.2) (38) and pseudotimes were calculated using Monocle3 (version 1.3.4) (39). TCR and BCR sequences were aligned separately in Cell Ranger and preprocessed using the scRepertoire pipeline (version 2.0.0) (40). Clonal information of TCR/BCR sequences was added to the Seurat object for downstream analysis. Interactome analysis was performed using the CellChat pipeline (version 2.1.2) (41).

### Cytometry by time of flight (CyTOF)

PBMC were thawed for bulk analysis in three independent experiments. PBMC were counted and 1×10^6^ cells were stained with metal-conjugated antibodies (Table S1) in Cell Staining Buffer (CSB; Standard BioTools, San Francisco, CA) for 30 minutes (surface stain) and 45 minutes (intracellular stain) at 4°C. Cells were washed, fixed with 1.6% PFA in PBS, subsequently pelleted, and resuspended in CSB (1×10^6^ cells per mL) for the acquisition on a CyTOF-XT instrument (Standard BioTools, San Francisco, CA) at the Longwood Medical Area CyTOF Core at the Dana Farber Cancer Institute. Raw data were normalized and debarcoded by the Mass Cytometry Core and live single nucleated cells were identified using Ir191/193 and Rh103. CD45 expression was evaluated and gated using FlowJo (version 10.9.0; BD Biosciences, Franklin Lakes, NJ), and CD45^+^ cells were imported into R (version 4.3.3) using R Studio (version 2023.12.0). Data were analyzed in a modified standard workflow (42) using FlowSOM (version 2.10.0) (43), CATALYST (version 1.26.1) (44), and flowCore (version 2.14.2) (45). The optimal number of meta clusters were determined using the Clustree method (version 0.5.1) (34).

### Serological IgG antibody concentrations

IgG antibody concentrations against the SARS-CoV-2 S1, S2, RBD, and nucleocapsid antigens were measured using the SARS-CoV-2 Antigen Panel 1 IgG (MilliporeSigma, Darmstadt, Germany). Samples were run in duplicate, and analysis was performed on a Luminex LX200 platform equipped with xPonent software (version 3.1.871.0; Luminex, Austin, Tx). Performance of the assay was validated using a clinically certified assay (Figure S6 and data not shown).

### Serological antibody neutralization assays

SARS-CoV-2 variant neutralization was assessed in a bead-based ACE2 competition assay (Bio-Plex Pro Human SARS-CoV-2 Neutralization Antibody Assay; Bio-Rad Laboratories, Hercules, CA). Ancestral and Delta S1-covered beads were purchased, and Omicron S1-covered beads were generated using the Custom Assay Developer Kit. Briefly, 1 µg Omicron RBD (Sino Biologics, Beijing, China) was coupled to magnetic COOH beads according to manufacturer’s instructions and assay performance was validated using plasma from three adult volunteers two weeks following SARS-CoV-2 infection as positive control and plasma from unvaccinated pediatric volunteers as negative control (data not shown).

### ELISPOT assays

Enzyme-linked Immuno Spot (ELISPOT) assays to measure IFN-γ production were performed on PMBCs collected from research subjects according to manufacturer’s instructions (Thermo Fisher Scientific, Waltham, MA). Briefly, CD4^+^ T cells were isolated using magnetic beads (Stemcell Technologies, Vancouver, Canada) and cultured in RPMI 1640 (Lonza, Walkersville, MD) supplemented with 10% FBS (Millipore Sigma, Darmstadt, Germany), 2 mM L-glutamine, 1 mM sodium pyruvate, 0.75 g/l sodium bicarbonate, 100 U/ml penicillin/streptomycin, 0.1 mM non-essential amino acids (all Lonza, Walkersville, MD) and 50 µM 2-mercaptoethanol (Millipore Sigma, Darmstadt, Germany) in 96-well polyvinylidene fluoride plates (Immobilon-P, Millipore, Billerica, MA). To assess cellular immunity against SARS-CoV-2, irradiated (1700 rads) PBMC from the same blood donor were added in a 1:1 ratio with CD4^+^ T cell responders in the presence of 10 µg/ml of either ancestral (Biolegend, San Diego, CA), Delta or Omicron S1 (both Sino Biologics, Beijing, China) peptide for 48 hours. Treatment of T cells with phytohaemagglutinin (PHA, 3 µg/ml) was used as a technical positive control. After staining, the plates were scanned and analyzed on an ImmunoSpot S6 Ultra ELISpot reader (version 5.0, CTL, Shaker Heights, OH).

### Statistics

Statistical tests were performed using the two-tailed Student’s t-test or One-way or Two-way ANOVA as indicated with previous testing of equality of variances. The Mann-Whitney test or Kruskal-Wallis test were used if variances were significantly different. As indicated, an appropriate post-hoc test with correction for multiple testing was performed to identify exactly which groups differ from each other. Multiple Mann-Whitney testing was corrected using the two-stage linear step-up procedure of Benjamini, Krieger and Yekutieli (46) and corrected *P* values are called *Q* values. Correlations between two cohorts were identified using Spearman’s rank correlation coefficient (Rho) and *P* values were calculated using algorithm AS 89 (47). *P* and *Q* values of less than 0.05 were considered significant.

### Study approval

The study and protocols were approved by the Institutional Review Board of the Boston Children’s Hospital. Written informed consent was received prior to participation.

### Data availability

Data reported in this paper will be shared by the corresponding authors upon request. RNA-seq data have been deposited at the National Center for Biotechnology Information Gene Expression Omnibus as GSE282427 and are publicly available as of the date of publication. Any additional information required to reanalyze the data reported in this paper is available from the corresponding authors upon request.

## Author contributions

The 2 first coauthors, JW and YT, each made unique and critical contributions to this manuscript and agreed on the order of authorship based on the lead taken in coordinating manuscript development. Conceptualization: JW, YT, KDM, LSK, SBS, DMB, BHH; Experimental design: JW, YT, MAS, LSK, SBS, DMB, BHH; Patient recruitment and consent: JW, LVC, MN, IO, KW, NWL, DMB; Sample collection: JW, BA, MM, LVC, MB, GS, RB; Methodology, Resources and Assay development: JW, YT, BS, MM, NJC; Performed experiments: JW, YT, BA, MM, RF, MAS, UG, AA; Data analysis: JW, YT, VD, BHH; Data interpretation: JW, YT, SBS, DMB, BHH; Scientific discussions: JW, YT, MS, UG, AA, VM, LVC, SJS, FW, KDM, LSK, SBS, DMB, BHH; Writing - original draft: JW, BHH; Writing - review and editing: JW, YT, DMB, BHH; Acquisition of funding: SBS, DMB, LSK, KDM.

## Acknowledgments

We thank Nancy Rodig, MD, Kevin P. Daly, MD, Scott A. Elisofon, MD, Emily Toal, Brendan Kimball and Morgan S Miovski for help with patient recruitment, sample collection and processing, Sek Won Kong, MD for helpful scientific discussions, and Alvin Kho, PhD for statistical support (all Boston Children’s Hospital). This study was supported by a Clusters of Clinical Research Excellence Award from Boston Children’s Hospital to S.B.S, D.M.B, L.S.K and K.D.M..

