## Supplemental Material for "Personalized Immune Profiling in Pediatric Transplant Recipients: Linking Atypical B Cells to Vaccine Response"

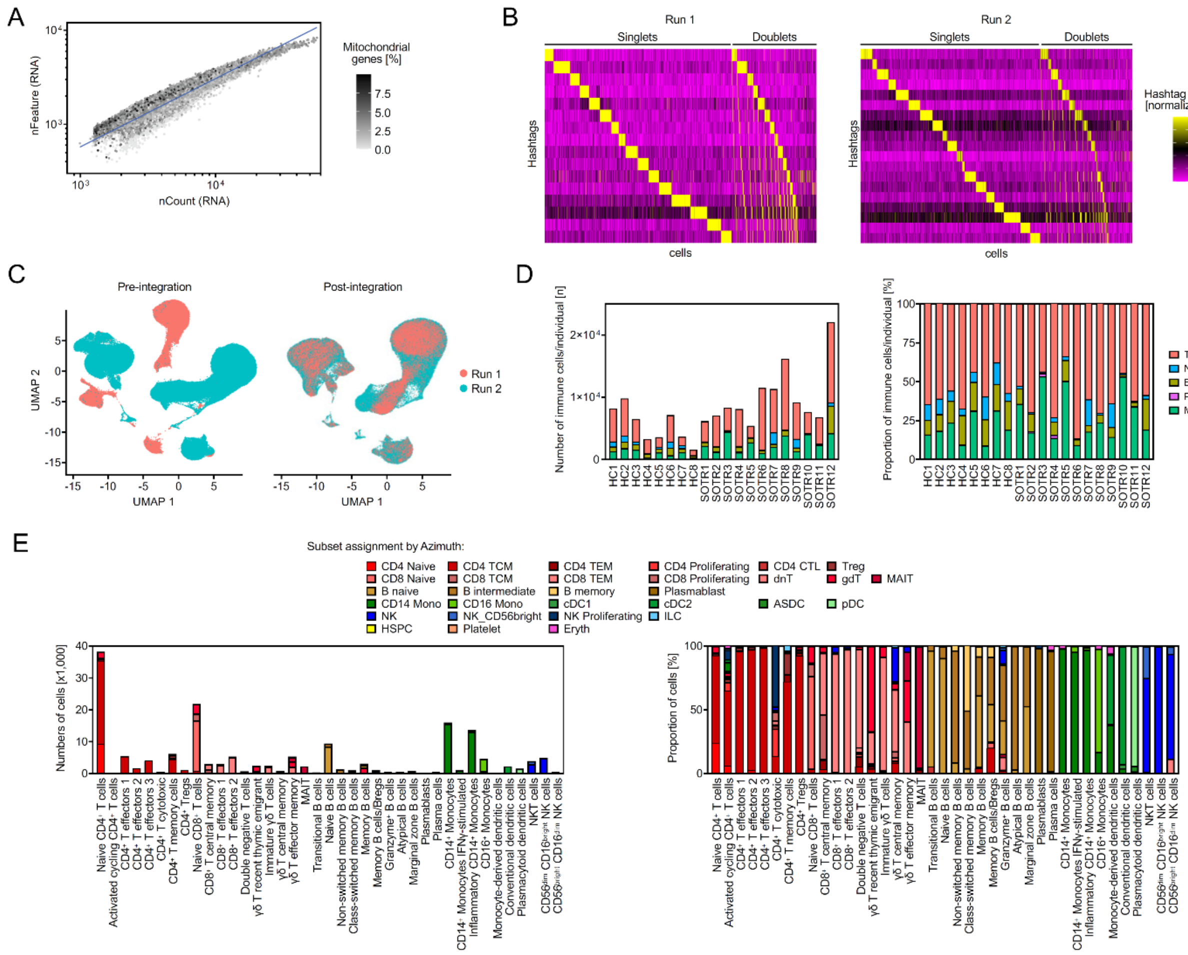


**Figure S1: QC matrices of scRNAseq experiment.**
**(A)** Scatter plot depicts counts of unique genes (nFeature) versus the total number of detected genes per cell post-QC. The color represents the percentage of mitochondrial genes among all genes per cell. **(B)** Samples were demultiplexed using hashtag antibodies. Heatmaps depict normalized hashtag expression of 5,000 randomly selected cells for each of the two independent sequencing runs. Cells with one hashtag were considered singlets, while cells with multiple hashtags were regarded as doublets and excluded from the analysis. **(C)** Batches of independent sequencing runs were corrected using Harmony [33]. UMAPs depict cells prior (left) and following successful integration (right). **(D)** Stacked bar graphs illustrate the absolute number (left) and proportion (right) of immune cell populations per subject. **(E)** Cell subset annotation was performed manually (x-axis) and subsets were validated using the Azimuth cell annotation tool. Bar graphs illustrate the absolute (left) and proportion (right) of automated recognized cells per manually annotated cell subset.

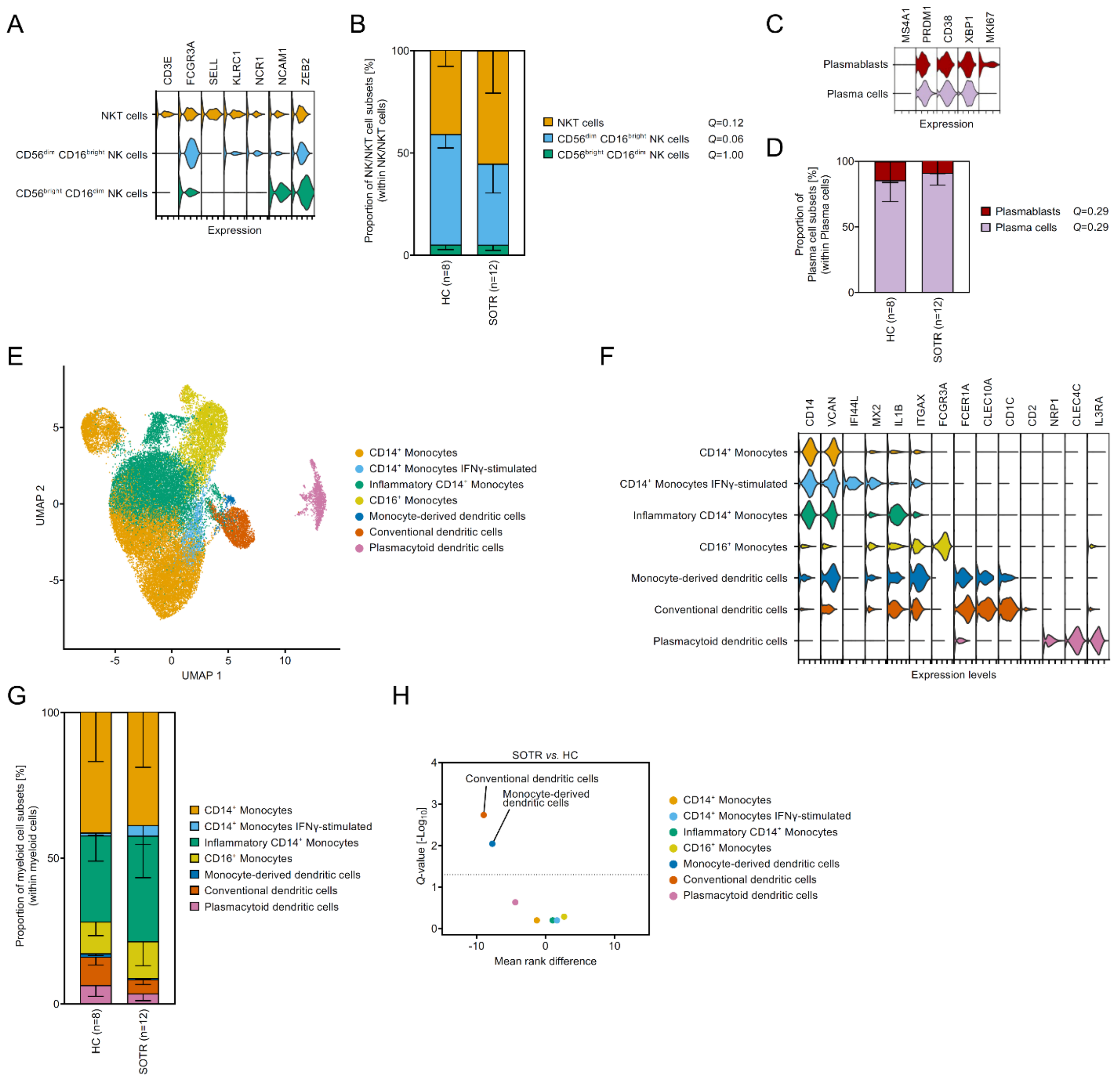


**Figure S2: Subclustering and quantification of NK/NKT, plasma and myeloid cell subsets.** NK, NKT **(A-B)**, plasma **(C-D)** and myeloid **(E-H)** cells were subclustered and the frequencies of cell subsets and phenotypes were evaluated. **(A)** Violin plot illustrates markers that were used to identify NK/NKT cell subsets. **(B)** Stacked bar graph represents NK/NKT cell subcluster distribution in SOTR *vs.* HC (mean ± SD). The corrected *Q*-value of NK/NKT cell subset frequencies in SOTR *vs.* HC was calculated using the Mann-Whitney test with two-stage linear step-up procedure of Benjamini, Krieger and Yekutieli. **(C)** Violin plot illustrates markers that were used to identify Plasma cell subsets. **(D)** Stacked bar graph represents plasmablast and plasma cell distribution in SOTR *vs.* HC (mean ± SD). The corrected *Q*-value of Plasma cell subset frequencies in SOTR *vs.* HC was calculated using the Mann-Whitney test with two-stage linear step-up procedure of Benjamini, Krieger and Yekutieli. **(E)** UMAP of 39,262 myeloid cells from 20 participants (8 healthy controls and 12 SOTR) color-coded for myeloid cell subsets. **(F)** Violin plot illustrates markers that were used to identify myeloid subsets. **(G)** Stacked bar graph represents myeloid subcluster distribution in SOTR *vs.* HC (mean ± SD). **(H)** Volcano plot depicts the mean rank difference and the corrected *Q*-value of myeloid subset frequencies in SOTR *vs.* HC (Mann-Whitney test with two-stage linear step-up procedure of Benjamini, Krieger and Yekutieli)**.**

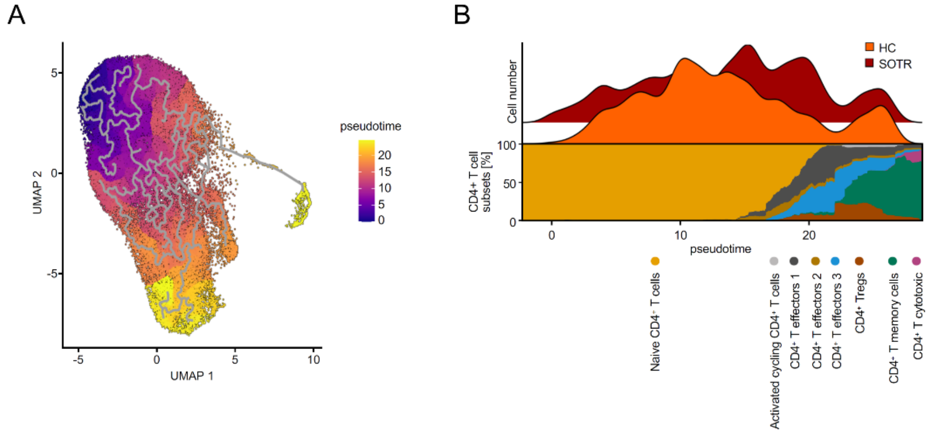


**Figure S3: Pseudotime trajectory analysis of CD4$^+$ T cell subsets in SOTR *vs.* HC.**
**(A)** UMAP color-coded for calculated pseudotime. **(B)** Ridge plot (top) and cluster heatmap (bottom) depict the frequencies of cells along a pseudotime.

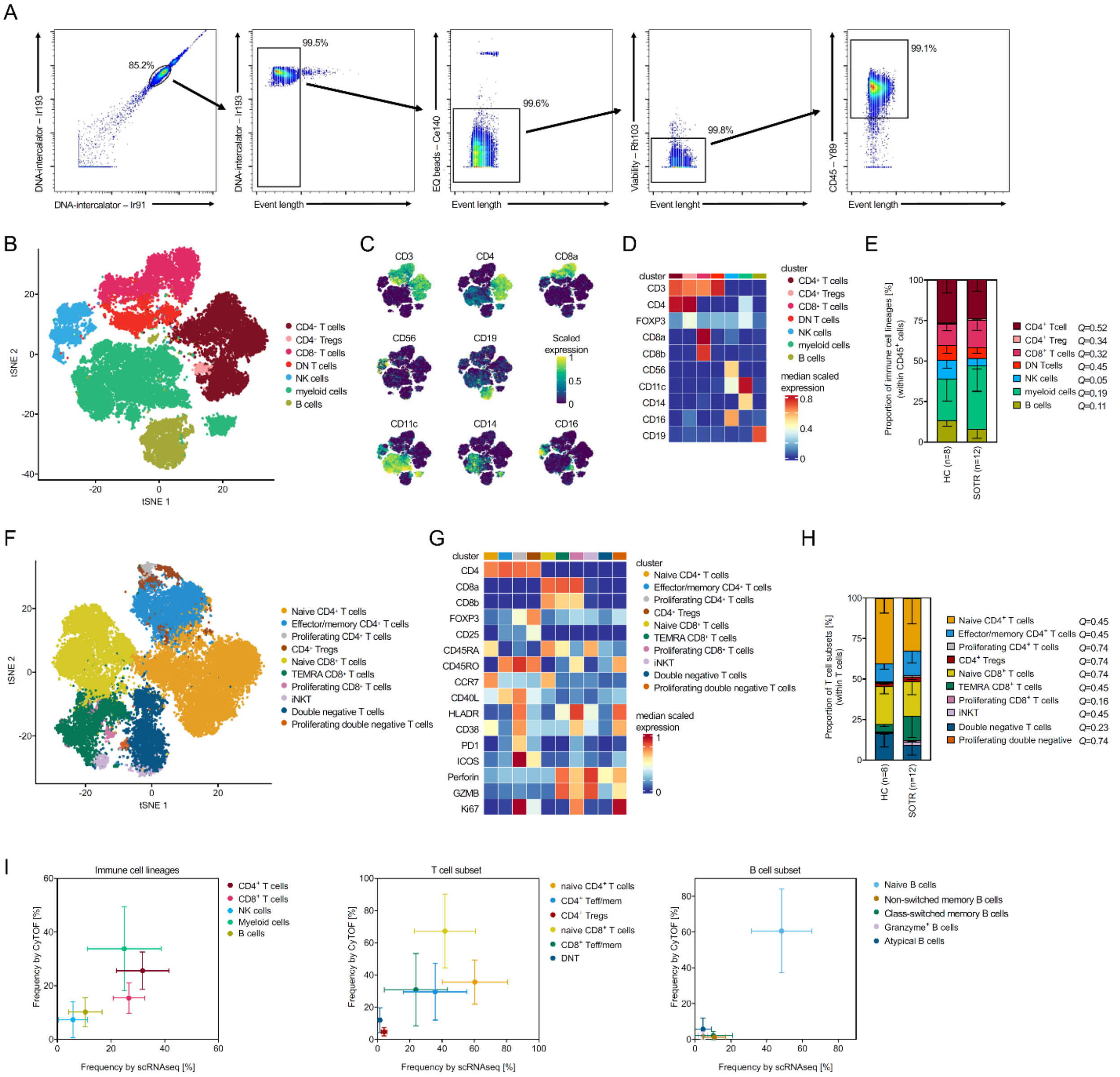


**Figure S4: Coarse clustering of PBMC using CyTOF validates no major differences in immune lineage populations between pediatric SOTR and healthy children.**

PBMC were isolated from blood samples, stained with immune cell lineage marker antibodies (see also Table S2) and CyTOF was performed to identify immune cell frequencies. **(A)** Dot plots illustrate the gating strategy to identify single viable CD45$^{pos}$ cells prior to import into R for analysis. **(B)** tSNE scatter plot depicts CD45$^{pos}$ PBMC color-coded by immune cell lineage. **(C)** Feature plots illustrate lineage marker expression used to identify immune lineages. **(D)** Heatmap depicts the expression of immune cell lineage defining markers. **(E)** Stacked bar graph illustrates frequencies of major cell lineages per subject and cohort (mean ± SD). The corrected *Q*-value comparing the same immune lineage between SOTR and HC was calculated using the Mann-Whitney test with two-stage linear step-up procedure of Benjamini, Krieger and Yekutieli. **(F)** tSNE plot depicts subclustered CD3$^{+}$ T cells color-coded for identified subsets. **(G)** Heatmap illustrates the expression of T cell subset defining markers. **(H)** Stacked bar graph illustrates frequencies of T cell subsets in SOTR *vs.* HC (mean ± SD; Mann-Whitney test with two-stage linear step-up procedure of Benjamini, Krieger and Yekutieli). **(I)** Scatter plot illustrates the correlation between subset frequencies of scRNA-seq and CyTOF analyses.

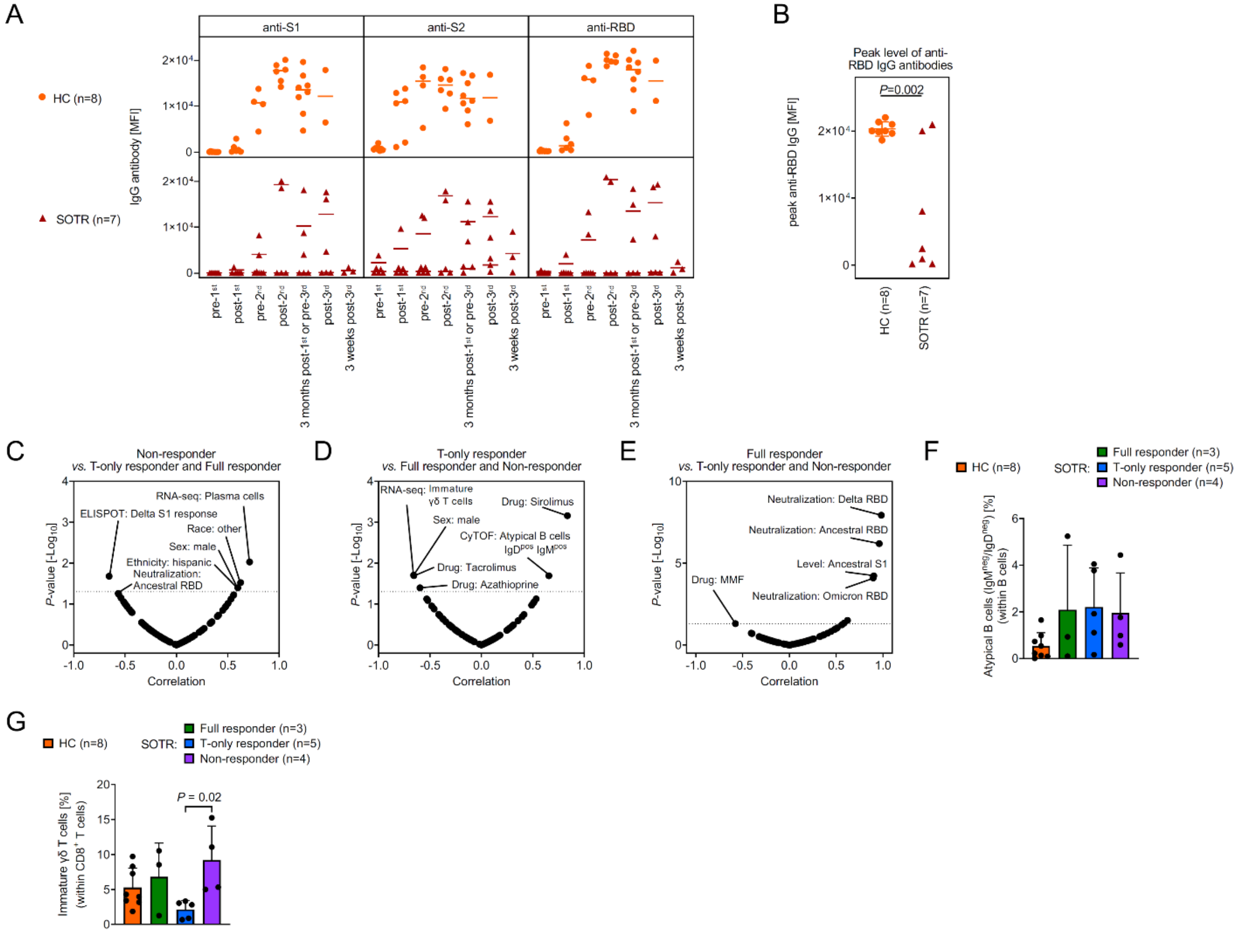


**Figure S5: Correlation analysis to identify associations with impaired protective immunity in pediatric SOTR.**

**(A)** Serological antibody levels of anti-S1, anti-S2 and anti-receptor-binding domain (RBD) of SARS-CoV-2 in pediatric SOTR and healthy children following immunizations with the BNT162b2 vaccine. **(B)** Ancestral anti-RBD IgG peak levels of HC and SOTR were compared using the Kruskal-Wallis test. **(C-E)** Correlations of all demographic data, frequencies of cell subsets as assessed by scRNAseq and CyTOF analyses, and SARS-CoV-2 antibody serum concentrations, neutralization capacity and ELISPOT response with either non-responsiveness **(C)**, T-responsiveness **(D)** or full responsiveness **(E)** to BNT162b2 vaccination were calculated using the Spearman's rank correlation test. Scatter plot depicts the Spearman's correlation coefficient versus the *P*-value. The dotted line indicates the significance level $P$=0.05. **(F)** Bar graph compares frequencies of IgM$^{neg}$/IgD$^{neg}$ atypical B cells between HC and SOTR subgroups (Kruskal-Wallis test with Dunn's multiple comparison; all comparisons are $P$>0.05). **(G)** Bar graph compares frequencies of Immature γδ T cells between HC and SOTR subgroups (Kruskal-Wallis test with Dunn's multiple comparison).

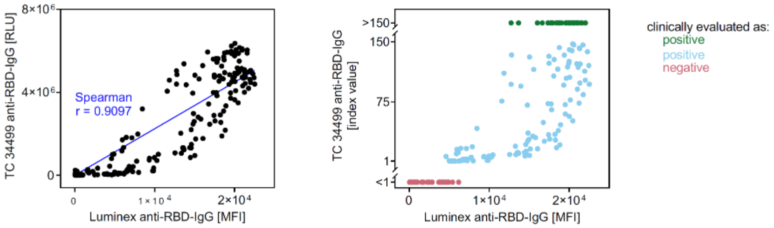


**Figure S6: Validation of Luminex assay.**

**(A)** A subset of serum samples was analyzed using the multiplexed bead ELISA (Luminex) and a semiquantitative commercial anti-S1 IgG assay (TC34499). Scatter plots depict the comparison of both assays. ***Left:*** Raw values (relative light units) of TC34499 were compared to our assay. The Spearman's rank correlation coefficient was calculated. ***Right:*** Clinical reported index units of TC34499 were compared to our data to illustrate sensitivity of our analysis.

| Metal conjugation | Reagent | Manufacturer | Clone | Catalog No. |
|---|---|---|---|---|
| 103Rh | cationic nucleic acid intercalator | STB | N/A | 201103A |
| 191/193Ir | cationic nucleic acid intercalator | STB | N/A | 201192A |
| 111Cd | anti-human Tbet Antibody | Core | 4B10 | N/A |
| 112Cd | anti-human CD8a Antibody | Core | RPA-T8 | N/A |
| 114Cd | anti-human CD4 Antibody | Core | RPA-T4 | N/A |
| 115In | anti-human CD3 Antibody | Core | UCHT1 | N/A |
| 116Cd | anti-human CD56 Antibody | Core | NCAM16.2 | N/A |
| 141Pr | anti-human CD196 (CCR6) Antibody | Core | G034E3 | N/A |
| 142Nd | anti-human CD19 Antibody | Core | HIB19 | N/A |
| 143Nd | anti-human CD127 Antibody | Core | eBioRDR5 | N/A |
| 144Nd | anti-human CD14 Antibody | Core | M5E2 | N/A |
| 145Nd | anti-human CD8b Antibody | Core | SIDI8BEE | N/A |
| 146Nd | anti-human IgD Antibody | STB | IA6-2 | 3146005B |
| 147Sm | anti-human CD45RO Antibody | Core | REA611 | N/A |
| 148Nd | anti-human CD69 Antibody | Core | FN50 | N/A |
| 149Sm | anti-human CD25 Antibody | Core | M-A251 | N/A |
| 150Nd | anti-human CD134 (OX40) Antibody | STB | ACT35 | 3150023B |
| 151Eu | anti-human IgG Antibody | Core | G18-145 | N/A |
| 152Sm | anti-human CD21 Antibody | Core | Bu32 | N/A |
| 153Eu | anti-human CD194 (CCR4) Antibody | Core | L291H4 | N/A |
| 154Sm | anti-human CD38 Antibody | Core | HIT2 | N/A |
| 155Gd | anti-human CD279 (PD-1) Antibody | STB | EH12.2H7 | 3155009B |
| 156Gd | Recombinant SARS-CoV-2 S1 Protein | BioLegend | N/A | 792904 |
| 158Gd | anti-human CD137 (4-1BB) Antibody | STB | 4B4-1 | 3158013B |
| 159Tb | anti-human CD11c Antibody | Core | Bu15 | N/A |
| 160Gd | anti-human CD278 (ICOS) Antibody | Core | C398.4A | N/A |
| 161Dy | anti-human CD138 Antibody | Core | MI15 | N/A |
| 162Dy | anti-human CD16 Antibody | Core | 3G8 | N/A |
| 163Dy | anti-human CD183 (CXCR3) Antibody | STB | G025H7 | 3163004B |
| 164Dy | anti-human Ki67 Antibody | Core | 8D5 | N/A |
| 165Ho | anti-human FoxP3 Antibody | Core | PCH101 | N/A |
| 166Er | anti-human CD24 Antibody | STB | ML5 | 3166007B |
| 167Er | anti-human CD27 Antibody | STB | L128 | 3167006B |
| 168Er | anti-human CD154 (CD40L) Antibody | STB | 24-31 | 3168006B |
| 169Tm | anti-human CD95 (FAS) Antibody | Core | DX2 | N/A |
| 170Er | anti-human CD45RA Antibody | STB | HI100 | 3170010B |
| 171Yb | anti-human CD185 (CXCR5) Antibody | STB | RF8B2 | 3171014B |
| 172Yb | anti-human IgM Antibody | Core | MHM-88 | N/A |
| 173Yb | anti-human GZMB Antibody | STB | GB11 | 3173006B |

| | | | | |
|---|---|---|---|---|
| 174Yb | anti-human HLA-DR Antibody | Core | L243 | N/A |
| 175Lu | anti-human Perforin Antibody | STB | B-D48 | 3175004B |
| 176Yb | anti-human CD197 (CCR7) Antibody | Core | G043H7 | N/A |
| 89Y | anti-human CD45 Antibody | STB | HI30 | 3089003B |

**Table S1. List of metal-conjugated antibodies for CyTOF.** STB: Standard BioTools. Core: Harvard Medical Area CyTOF Antibody Resource and Core (Lederer Lab at Brigham and Women's Hospital) https://ledererlab.bwh.harvard.edu/cytof-core/

| DNA barcode | Reagent | Manufacturer | Clone | Catalog No. |
|---|---|---|---|---|
| GTCAACTCTTTAGCG | TotalSeq-C0251 anti-human Hashtag 1 Antibody | BioLegend | LNH-94 and 2M2 | 394661 |
| TGATGGCCTATTGGG | TotalSeq-C0252 anti-human Hashtag 2 Antibody | BioLegend | LNH-94 and 2M2 | 394663 |
| TTCCGCCTCTCTTTG | TotalSeq-C0253 anti-human Hashtag 3 Antibody | BioLegend | LNH-94 and 2M2 | 394665 |
| AGTAAGTTCAGCGTA | TotalSeq-C0254 anti-human Hashtag 4 Antibody | BioLegend | LNH-94 and 2M2 | 394667 |
| AAGTATCGTTTCGCA | TotalSeq-C0255 anti-human Hashtag 5 Antibody | BioLegend | LNH-94 and 2M2 | 394669 |
| GGTTGCCAGATGTCA | TotalSeq-C0256 anti-human Hashtag 6 Antibody | BioLegend | LNH-94 and 2M2 | 394671 |
| TGTCTTTCCTGCCAG | TotalSeq-C0257 anti-human Hashtag 7 Antibody | BioLegend | LNH-94 and 2M2 | 394673 |
| CTCCTCTGCAATTAC | TotalSeq-C0258 anti-human Hashtag 8 Antibody | BioLegend | LNH-94 and 2M2 | 394675 |
| CAGTAGTCACGGTCA | TotalSeq-C0259 anti-human Hashtag 9 Antibody | BioLegend | LNH-94 and 2M2 | 394677 |
| ATTGACCCGCGTTAG | TotalSeq-C0260 anti-human Hashtag 10 Antibody | BioLegend | LNH-94 and 2M2 | 394679 |
| TAACGACCAGCCATA | TotalSeq-C0262 anti-human Hashtag 12 Antibody | BioLegend | LNH-94 and 2M2 | 394683 |
| AAATCTCTCAGGCTC | TotalSeq-C0263 anti-human Hashtag 13 Antibody | BioLegend | LNH-94 and 2M2 | 394685 |
| CTGTATGTCCGATTG | TotalSeq-C0264 anti-human Hashtag 14 Antibody | BioLegend | LNH-94 and 2M2 | 394687 |
| TAAGATTCAGAGCGA | TotalSeq-C0265 anti-human Hashtag 15 Antibody | BioLegend | LNH-94 and 2M2 | 394689 |
| CTCAGTGCATTCTGG | TotalSeq-C0276 anti-human Hashtag 16 Antibody | BioLegend | LNH-94 and 2M2 | 394691 |
| CAAACCAACAGTTCG | TotalSeq-C0278 anti-human Hashtag 18 Antibody | BioLegend | LNH-94 and 2M2 | 394693 |
| ACCCTTCCCTTCGTT | TotalSeq-C0279 anti-human Hashtag 19 Antibody | BioLegend | LNH-94 and 2M2 | 394695 |
| TAACCACTGGATGAT | TotalSeq-C0280 anti-human Hashtag 20 Antibody | BioLegend | LNH-94 and 2M2 | 394697 |
| GGTTGTTGTTAGTGG | TotalSeq-C0284 anti-human Hashtag 24 Antibody | BioLegend | LNH-94 and 2M2 | 394699 |

**Table S2. List of Hashtag antibodies.**
